# Repurposing Ivermectin and ATRA as Potential Therapeutics for Glioblastoma Multiforme

**DOI:** 10.1101/2024.08.26.24312575

**Authors:** Saed Sayad, Mark Hiatt, Hazem Mustafa

## Abstract

**Background:** Glioblastoma multiforme (GBM) is the most aggressive and lethal form of primary brain tumor, characterized by rapid growth and resistance to conventional therapies. Despite advances in treatment, most patients succumb to the disease within 15 months. Drug repurposing, which involves finding new uses for existing drugs, is a promising strategy to develop new GBM treatments faster and more cost-effectively.

**Method:** We obtained single-cell RNA sequencing (scRNA-seq) data (*<u>GSE84465</u>*) from the National Institutes of Health (NIH) Gene Expression Omnibus (GEO) repository to compare gene expression in GBM neoplastic cells and non-neoplastic cells. We identified genes that were abnormally expressed in tumor cells and linked these genes to potential drug targets. To identify potential repurposed drugs for GBM, we leveraged the Chemical Entities of Biological Interest (ChEBI) database to assess the interaction of various compounds with the differentially expressed genes identified in the scRNA-seq analysis. We focused on compounds that could reverse the aberrant gene expression observed in GBM neoplastic cells.

**Results:** Our analysis suggests that ivermectin and all-trans-retinoic acid (ATRA) could be repurposed as effective treatments for GBM. Ivermectin, typically used as an antiparasitic, demonstrated strong anti-tumor activity by downregulating 40 of the top 100 upregulated genes in GBM, indicating its potential to suppress tumor growth. ATRA, known for promoting cell differentiation, upregulated 60 genes typically downregulated in GBM neoplastic cells, showing its potential to correct transcriptional dysregulation and support tumor suppression. These findings underscore the promise of drug repurposing to target key pathways in GBM, offering new therapeutic options for this aggressive cancer.

**Conclusions:** Our results provide compelling evidence that ivermectin and ATRA may be effective in treating GBM. The observed alterations in gene expression indicate the ability of these two agents to disrupt key genes and pathways crucial for tumor progression. Given the increasing interest in drug repurposing for cancer treatment, comprehensive preclinical and clinical investigations are warranted to assess fully the therapeutic efficacy of these compounds against this disease.

## Introduction

Glioblastoma multiforme (GBM) is the most aggressive and lethal form of primary brain tumor, characterized by rapid growth, extensive invasion into surrounding brain tissue, and a highly heterogeneous cellular and molecular landscape. Despite advances in surgical resection, radiation therapy, and chemotherapy, the prognosis for GBM patients remains dismal, with a median survival of only 15 months. The standard treatments of radiation, chemotherapy, and surgery provide limited efficacy, and nearly all patients experience tumor recurrence. This dire outlook underscores the urgent need for novel therapeutic strategies. Drug repurposing, the strategy of identifying new therapeutic uses for existing drugs, has emerged as a promising approach to accelerate the development of treatments for GBM. Unlike traditional drug discovery, which can take over a decade and substantial financial investment to bring a new drug to market, repurposing leverages the known safety profiles, pharmacokinetics, and manufacturing processes of existing drugs. This approach not only reduces the time and cost of development, but also increases the likelihood of clinical success. Given the complexity of GBM biology, repurposing offers a practical way to identify agents that can modulate these pathways and improve outcomes. However, for GBM, the challenge remains to identify compounds that can effectively penetrate into the central nervous system, reach therapeutic concentrations within the tumor, and exert anti-tumor effects without significant toxicity.

## Data

We obtained scRNA-seq data sourced from the National Institutes of Health (NIH) Gene Expression Omnibus (GEO) repository (*<u>GSE84465</u>*). The dataset contains differential expression from cells within and in proximity of the tumor (Figures 1, 2, and 3).

**Figure 1:**
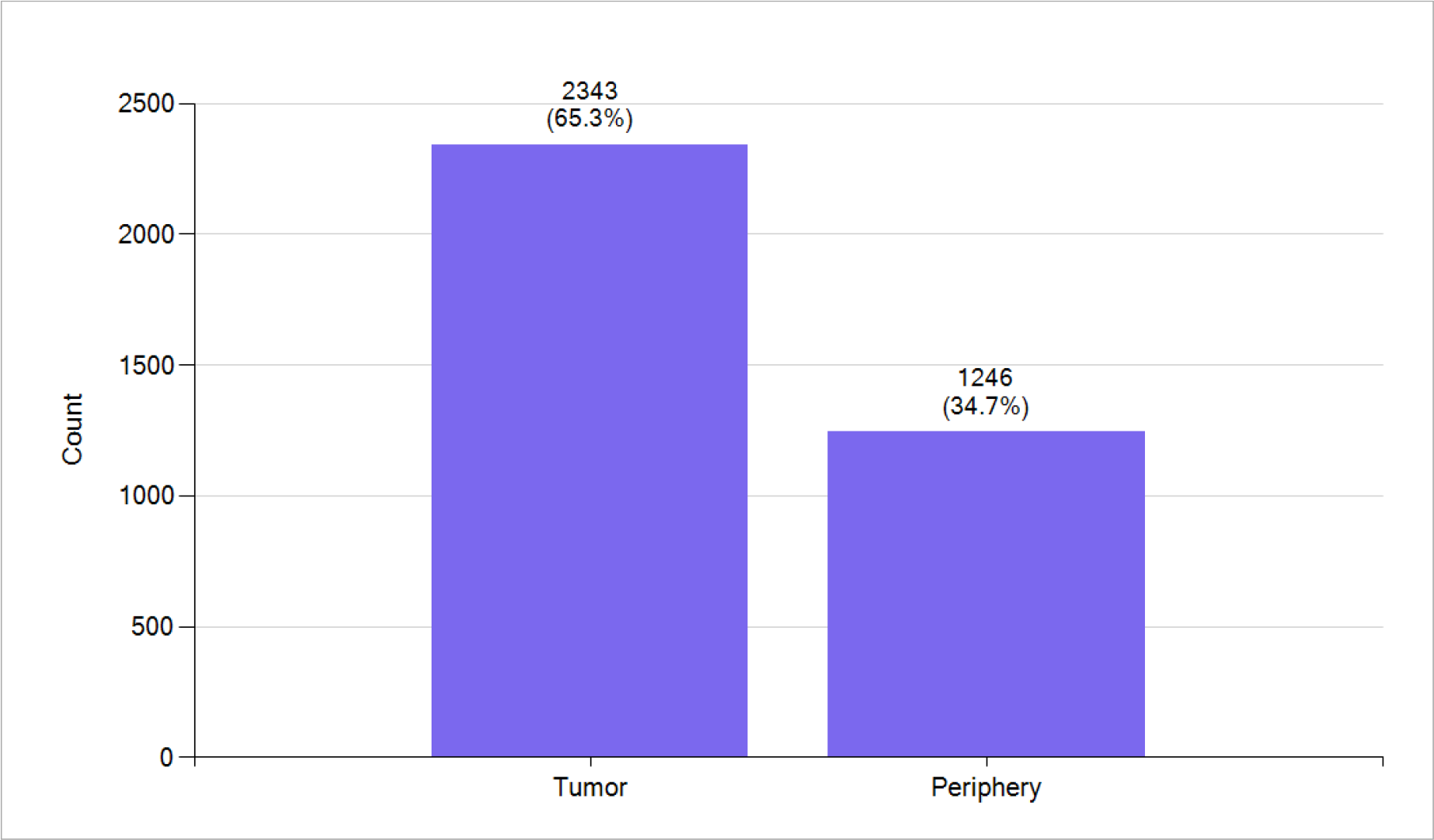
*<u>GSE84465</u>* contains 2343 GBM tumor single cells and 1246 periphery single cells.

**Figure 2:**
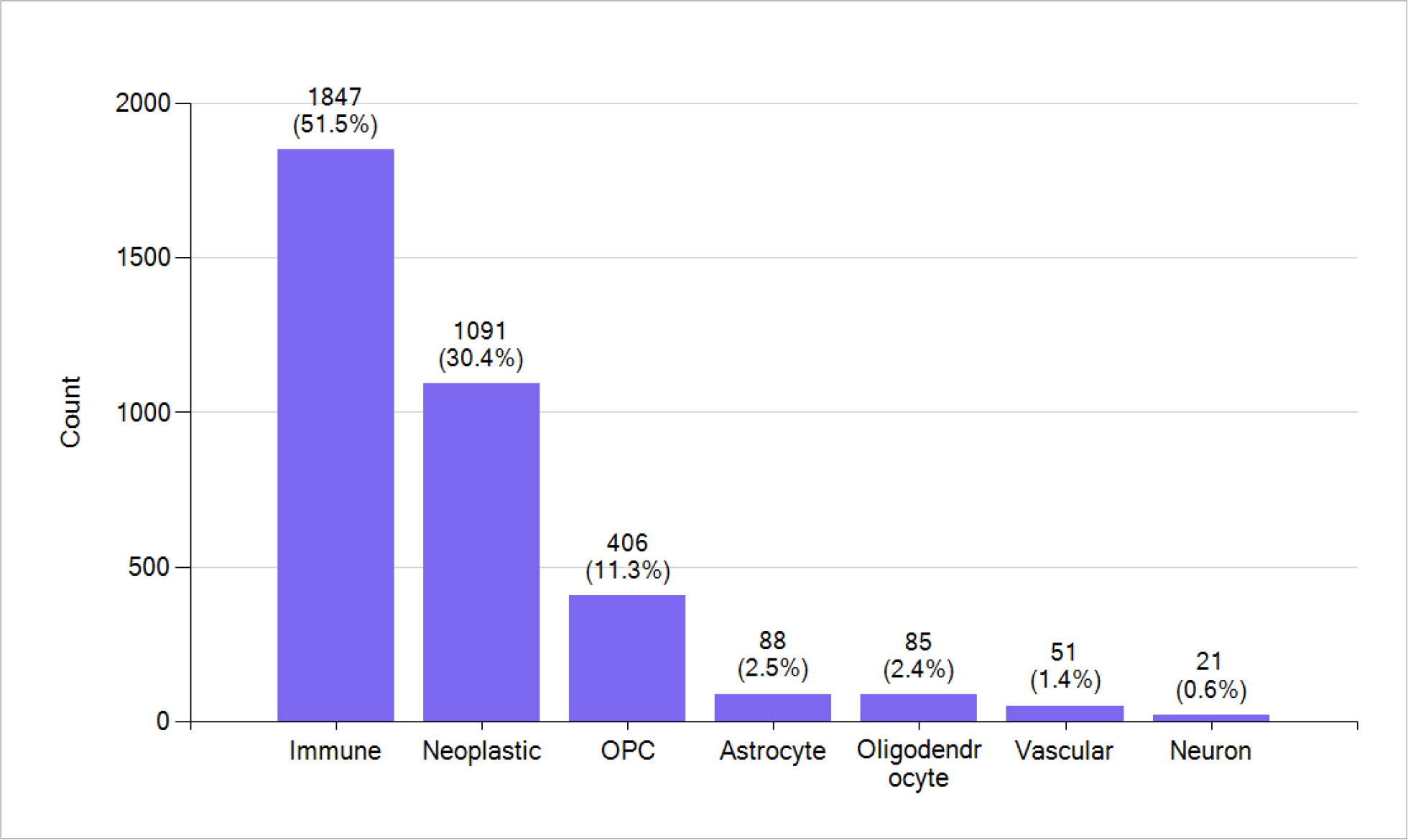
*<u>GSE84465</u>* includes seven different cell types. Oligodendrocyte progenitor cells (OPC) are a subtype of glia in the central nervous system named for their essential role as precursors to oligodendrocytes and myelin.

**Figure 3:**
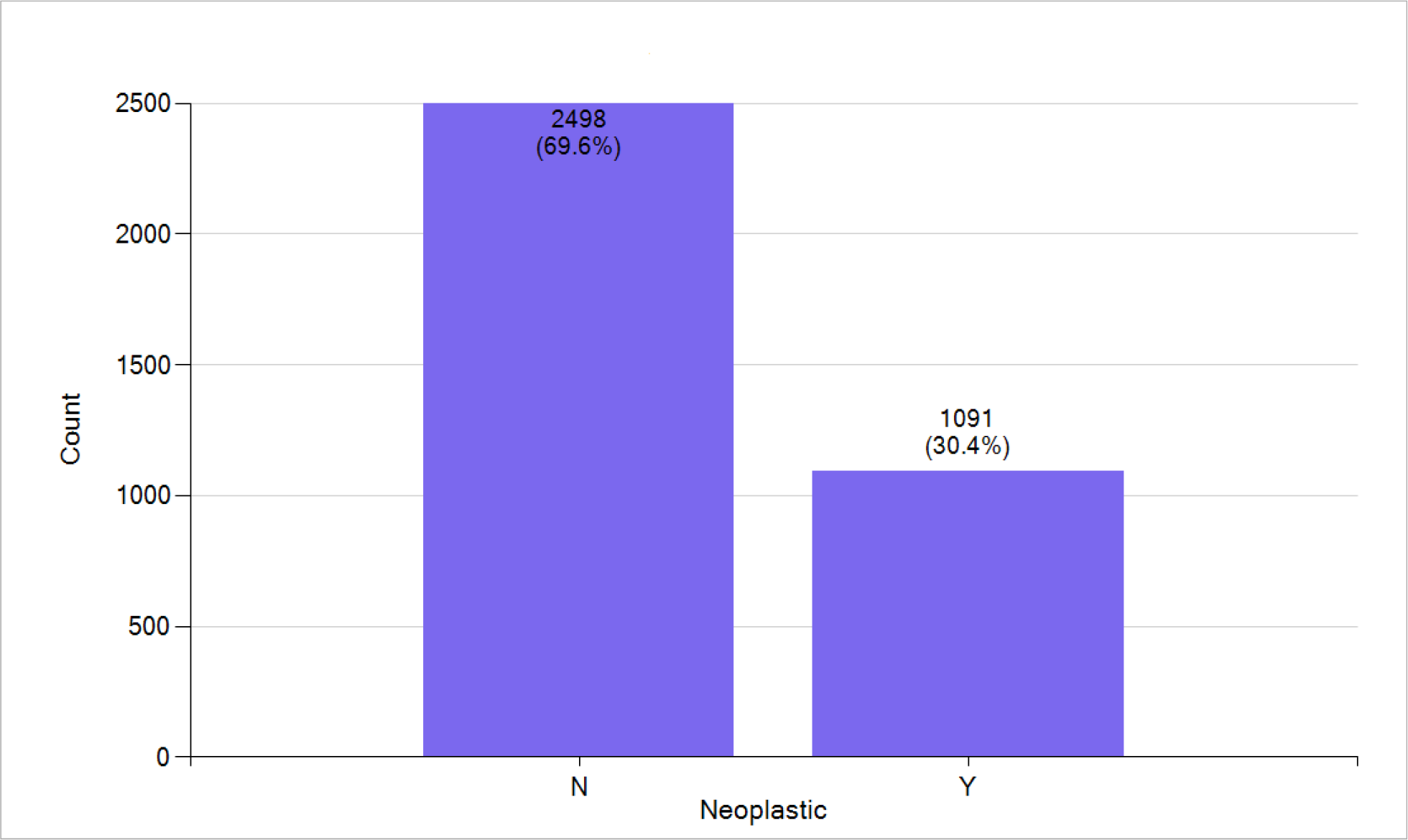
GBM non-neoplastic and neoplastic single cells.

### Differential gene expression – Non-neoplastic compared to neoplastic cells

Table 1 presents the top 10 genes that are either downregulated or upregulated when comparing non-neoplastic to neoplastic single cells. Downregulation and upregulation refer to the decrease or increase in gene expression, respectively.

**Table 1:** Top 10 downregulated and upregulated genes through single-cell gene analysis comparing non-neoplastic to neoplastic single cells.

| Non-neoplastic Compared to Neoplastic cells |  |  |  |
| --- | --- | --- | --- |
| Downregulated in Neoplastic cells |  | Upregulated in Neoplastic cells |  |
| Gene Symbol | Gene Name | Gene Symbol | Gene Name |
| *SRGN* | Serglycin | *CLU* | Clusterin |
| *LAPTM5* | Lysosomal Protein Transmembrane 5 | *PTPRZ1* | Protein Tyrosine Phosphatase Receptor Type Z1 |
| *TYROBP* | Transmembrane Immune Signaling Adaptor TYROBP | *CNN3* | Calponin 3 |
| *FCER1G* | Fc Epsilon Receptor Ig | *C1orf61* | MIR9-1 Host Gene |
| *CD74* | CD74 Molecule | *MT3* | Metallothionein 3 |
| *AIF1* | Allograft Inflammatory Factor 1 | *EGFR* | Epidermal Growth Factor Receptor |
| *C1QA* | Complement C1q A Chain | *PTN* | Pleiotrophin |
| *C1QB* | Complement C1q B Chain | *ITGB8* | Integrin Subunit Beta 8 |
| *IFI30* | IFI30 Lysosomal Thiol Reductase | *MAP1B* | Microtubule Associated Protein 1B |
| *SLC11A1* | Solute Carrier Family 11 Member 1 | *ATP1B2* | ATPase Na<sup>+</sup>/K<sup>+</sup> Transporting Subunit Beta 2 |

Table 1 lists genes that are downregulated in neoplastic cells compared to non-neoplastic ones. *S*erglycin, a proteoglycan involved in immune responses, works in an autocrine manner. Its suppression potently reduces their malignant properties, pro-neoplastic signaling, and stemness, and evokes astrocytic differentiation [1]. Lysosomal Protein Transmembrane 5 is associated with lysosomal function in cancer cells. Lysosomes undergo both qualitative and quantitative modifications that benefit the tumor. As a result, these organelles become promising targets for manipulating non-oncogene addiction [2]. Transmembrane Immune Signaling Adaptor TYROBP binds non-covalently to activated receptors on the surface of various immune cells, and mediates signal transduction and cellular activation. It is dysregulated in various malignancies [3]. Also downregulated is Fc Epsilon Receptor Ig (FCER1G), which acts as a key gene involved in cancer immune infiltration and the tumor microenvironment. Diminished cell proliferation promotes natural killer cell adaptive-like phenotype by limiting FCER1G expression [4]. CD74 Molecule, a type-II transmembrane glycoprotein associated with tumorigenesis, has been proposed as a prognostic marker and indicator of M1 macrophage infiltration in a comprehensive pan-cancer analysis [5]. Allograft Inflammatory Factor 1 encodes the tumor-associated-macrophage-specific protein IBA1, correlated with a worse prognosis in proneural GBM, but confers a survival benefit in mesenchymal tumors [6]. Additionally, the *C1QA* and *C1QB* components of the complement system, which are crucial for the immune response, are downregulated [7]. Other downregulated genes include IFI30 Lysosomal Thiol Reductase, which has been proposed as a novel immune-related target with predictive value in prognosis and treatment response in GBM [8], and Solute Carrier Family 11 Member 1, which plays a role in cellular resistance to pathogens and whose downregulation is associated with a favorable prognosis [9]. The downregulation of these genes suggests a potential impairment in immune function and antigen processing within neoplastic cells, possibly contributing to the immune evasion observed in GBM.

Conversely, the top 10 upregulated genes exhibit increased expression in neoplastic cells, indicating potential roles in tumor biology and progression. Clusterin is associated with various cellular processes, including stress response and apoptosis, and its expression is linked to carcinogenesis and prognosis [10]. Protein Tyrosine Phosphatase Receptor Type Z1 is involved in signaling pathways that may influence tumor growth. Its suppression has been shown to inhibit stem cell-like properties and tumorigenicity in glioblastoma [11]. Calponin 3 expression is increased in glioma, particularly GBM. Silencing expression of this gene inhibits the proliferation, migration, and invasion of glioma cell lines [12]. Metallothionein 3 is linked to metal ion regulation and high metallothionein predicts poor survival in GBM [13]. MIR9-1 Host Gene is associated with the regulation of microRNA, which plays a key role in many cellular processes, such as growth, differentiation, cell division, apoptosis, and cell signaling [14]. Epidermal Growth Factor Receptor, crucial for cell signaling, is frequently overexpressed in GBM, thus promoting growth and survival of neoplastic cells [15]. Pleiotrophin, involved in cell growth and angiogenesis, is a driver of vascular abnormalization in GBM [16]. Integrin Subunit Beta 8 is involved in angiogenesis. Pharmacological inhibition of this gene has been identified as a promising strategy for treating GBM [17]. Microtubule Associated Protein 1B contributes to microtubule dynamics and confers resistance to inhibition of mammalian target of rapamycin (mTOR) in human GBM [18]. Lastly, ATPase Na+/K+ Transporting Subunit Beta 2 is crucial for maintaining cellular ion balance and membrane potential. Elevated expression of of this gene has been linked to a significantly poorer prognosis in patients with GBM [19]. The upregulation of these genes suggests their involvement in tumor development and highlights potential targets for therapeutic intervention.

### Drug repurposing using compounds from the Chemical Entities of Biological Interest database

Drug repurposing, the strategy of identifying new therapeutic uses for existing drugs, is a promising approach in drug discovery that can significantly reduce the time and cost associated with bringing new treatments to market. The Chemical Entities of Biological Interest (ChEBI) database plays a vital role in this process by providing detailed information about small molecular entities. By leveraging ChEBI’s extensive and curated data, scientists can systematically search for drugs that interact with specific biological pathways or targets involved in diseases, thereby uncovering new therapeutic applications for existing compounds.

Table 2 shows the interaction of ChEBI compounds with the downregulated genes in the neoplastic cells of GBM. Among these, only two emerge as promising candidates for repurposing. The first, all-trans-retinoic acid (ATRA), can upregulate 60 genes that are downregulated in GBM while only downregulating four, highlighting its strong potential to activate gene expression and its promise as a therapeutic option for GBM. ATRA is known to induce cell differentiation and promote apoptosis in cancer cells, and its lipid-soluble nature allows it to easily cross the blood-brain barrier, making it particularly suitable for glioma treatment [20]. The second compound, acetamide, an organic molecule derived from acetic acid with the formula CH3CONH2, has practical uses as a plasticizer and industrial solvent. Notably, a novel acetamide-based HO-1 inhibitor has been shown to counteract GBM progression by interfering with the hypoxic-angiogenic pathway, further underscoring its potential as a therapeutic agent [21].

**Table 2:** Top 10 compounds that effectively upregulate/downregulate the top 100 downregulated genes in GBM cells. The compounds are listed alongside the number of genes they upregulate or downregulate.

| ChEBI Compounds and the Top 100 Downregulated Genes in GBM Cells |  |  |
| --- | --- | --- |
| Compound | Upregulated Genes by Compound | Downregulated Genes by Compound |
| all-trans-retinoic acid (ATRA) | 60 | 4 |
| acetamide | 56 | 6 |
| nickel atom | 54 | 5 |
| thioacetamide | 52 | 13 |
| carbon nanotube | 51 | 16 |
| 1,2-dimethylhydrazine | 46 | 10 |
| tetrachloromethane | 45 | 12 |
| titanium dioxide | 45 | 9 |
| lipopolysaccharide | 44 | 6 |
| cuprizon | 43 | 9 |

Table 3 presents the effects of various ChEBI compounds on the expression of the top 100 upregulated genes in GBM cells. Notably, ivermectin, while not upregulating any of the top 100 genes, downregulates 40 of them, indicating a significant suppressive effect on gene expression in GBM. It has been shown that ivermectin is effective in glioblastoma cells in vitro and in vivo by inhibiting angiogenesis, inducing mitochondrial dysfunction and oxidative stress, and deactivating the Akt/mTOR signaling pathway [22]. Other compounds such as sunitinib [23] and doxorubicin [24] also show considerable activity, upregulating 15 and 18 genes, respectively, while downregulating 36 and 31, respectively. The broad range of gene regulation observed across these compounds highlights their diverse molecular impacts on GBM cells, suggesting potential avenues for therapeutic intervention.

**Table 3:** Top 10 compounds that effectively downregulate the top 100 upregulated genes in GBM cells. The compounds are listed alongside the number of genes they upregulate or downregulate.

| ChEBI Compounds and the Top 100 Upregulated Genes in GBM Cells |  |  |
| --- | --- | --- |
| Compound | Upregulated Genes by Compound | Downregulated Genes by Compound |
| ivermectin | 0 | 40 |
| sunitinib | 15 | 36 |
| endosulfan | 9 | 33 |
| carbon nanotube | 21 | 32 |
| 6-propyl-2-thiouracil | 28 | 32 |
| doxorubicin | 18 | 31 |
| aristolochic acid A | 16 | 31 |
| paracetamol | 22 | 31 |
| acrylamide | 15 | 31 |
| folic acid | 6 | 29 |

The analysis of ChEBI compounds in Tables 2 and 3 underscores the potential of both ATRA and ivermectin as repurposed agents for the treatment of GBM. ATRA is distinguished by upregulating 60 genes that are typically downregulated in GBM while downregulating only four. This strong capacity to activate gene expression, coupled with its ability to induce cell differentiation, promote apoptosis, and cross the blood-brain barrier, positions ATRA as a promising candidate for GBM therapy. On the other hand, ivermectin, while not upregulating any of the top 100 genes, exhibits a remarkable ability to downregulate 40 of these genes, suggesting a potent suppressive effect on gene expression in GBM. Its known mechanisms of action, including the inhibition of angiogenesis, induction of mitochondrial dysfunction and oxidative stress, and deactivation of the Akt/mTOR signaling pathway, further support its potential as a therapeutic agent in GBM.

## Discussion

GBM remains one of the most formidable challenges in oncology, characterized by its aggressive behavior and resistance to conventional therapies. The inherent heterogeneity of GBM, driven by its complex cellular and molecular composition, necessitates innovative strategies to identify effective treatments. One such strategy is drug repurposing, which involves identifying new therapeutic uses for existing drugs. This approach is particularly appealing in the context of GBM due to the potential for rapid translation from bench to bedside. Our analysis of ChEBI compounds underscores the potential of both ivermectin and ATRA as repurposed agents for the treatment of GBM.

Ivermectin, traditionally used as an antiparasitic agent, demonstrates significant potential for repurposing in GBM treatment. Our analysis shows that ivermectin downregulates 40 of the top 100 upregulated genes in GBM, indicating a substantial suppressive effect on tumor gene expression. This compound’s ability to inhibit angiogenesis, induce mitochondrial dysfunction and oxidative stress, and deactivate the Akt/mTOR signaling pathway further supports its potential as an anti-tumor agent. These effects have been corroborated by studies demonstrating ivermectin’s efficacy in both in vitro and in vivo GBM models.

ATRA also emerges as a particularly promising candidate, with the ability to upregulate 60 genes that are typically downregulated in GBM, while downregulating only four. This activity suggests a strong potential to reverse the transcriptional dysregulation characteristic of GBM, particularly in pathways related to cell differentiation and apoptosis. ATRA’s known ability to induce differentiation and promote apoptosis in cancer cells, combined with its ability to cross the blood-brain barrier, underscores its suitability for glioma treatment.

The results of this study highlight the importance of exploring the transcriptional landscape of GBM at the single-cell level to uncover potential therapeutic targets and mechanisms of action. The ability of ivermectin and ATRA to modulate key pathways involved in GBM progression suggests that these compounds may offer novel therapeutic options for patients with this challenging disease. Furthermore, the use of existing drugs such as ivermectin and ATRA in a new therapeutic context could expedite the development of effective treatments for GBM, offering hope for improved outcomes in the future.

## Summary

In conclusion, our findings provide compelling evidence for the potential repurposing of ivermectin and ATRA as therapeutic agents for GBM. The significant alterations in gene expression observed in response to these compounds underscore their potential to disrupt key pathways involved in tumor growth and survival. As drug repurposing continues to gain traction as a viable strategy for cancer treatment, further preclinical and clinical investigations are warranted to fully elucidate the therapeutic potential of these compounds in GBM.

## Data Availability

All data produced are available online at:
https://www.ncbi.nlm.nih.gov/geo/query/acc.cgi?acc=GSE84465

## References

1. Tellez-Gabriel M, Tekpli X, Reine TM, Hegge B, Nielsen SR, Chen M, Moi L, Normann LS, Busund LR, Calin GA, Mælandsmo GM, Perander M, Theocharis AD, Kolset SO, Knutsen E. Serglycin Is Involved in TGF-β Induced Epithelial-Mesenchymal Transition and Is Highly Expressed by Immune Cells in Breast Cancer Tissue. Front Oncol. 2022 Apr 14;12:868868.

2. Jacobs KA, Maghe C, Gavard J. Lysosomes in glioblastoma: pump up the volume. Cell Cycle. 2020 Sep;19(17):2094–2104.

3. Lu J, Peng Y, Huang R, Feng Z, Fan Y, Wang H, Zeng Z, Ji Y, Wang Y, Wang Z. Elevated TYROBP expression predicts poor prognosis and high tumor immune infiltration in patients with low-grade glioma. BMC Cancer. 2021 Jun 23;21(1):723.

4. Yang R, Chen Z, Liang L, Ao S, Zhang J, Chang Z, Wang Z, Zhou Y, Duan X, Deng T. Fc Fragment of IgE Receptor Ig (FCER1G) acts as a key gene involved in cancer immune infiltration and tumour microenvironment. Immunology. 2023 Feb;168(2):302–319.

5. Li, R.Q., Yan, L., Zhang, L. et al. CD74 as a prognostic and M1 macrophage infiltration marker in a comprehensive pan-cancer analysis. Sci Rep 14, 8125 (2024).

6. Kaffes I, Szulzewsky F, Chen Z, Herting CJ, Gabanic B, Velázquez Vega JE, Shelton J, Switchenko JM, Ross JL, McSwain LF, Huse JT, Westermark B, Nelander S, Forsberg-Nilsson K, Uhrbom L, Maturi NP, Cimino PJ, Holland EC, Kettenmann H, Brennan CW, Brat DJ, Hambardzumyan D. Human Mesenchymal glioblastomas are characterized by an increased immune cell presence compared to Proneural and Classical tumors. Oncoimmunology. 2019 Aug 22;8(11):e1655360.

7. Bouwens van der Vlis TAM, Kros JM, Mustafa DAM, van Wijck RTA, Ackermans L, van Hagen PM, van der Spek PJ. The complement system in glioblastoma multiforme. Acta Neuropathol Commun. 2018 Sep 12;6(1):91.

8. Zhu C, Chen X, Guan G, Zou C, Guo Q, Cheng P, Cheng W, Wu A. IFI30 Is a Novel Immune-Related Target with Predicting Value of Prognosis and Treatment Response in Glioblastoma. Onco Targets Ther. 2020 Feb 5;13:1129–1143.

9. Takashima Y, Kawaguchi A, Kanayama T, Hayano A, Yamanaka R. Correlation between lower balance of Th2 helper T-cells and expression of PD-L1/PD-1 axis genes enables prognostic prediction in patients with glioblastoma. Oncotarget. 2018 Apr 10;9(27):19065–19078.

10. Fu Y, Du Q, Cui T, Lu Y, Niu G. A pan-cancer analysis reveals role of clusterin (CLU) in carcinogenesis and prognosis of human tumors. Front Genet. 2023 Jan 4;13:1056184.

11. Fujikawa A, Sugawara H, Tanaka T, Matsumoto M, Kuboyama K, Suzuki R, Tanga N, Ogata A, Masumura M, Noda M. Targeting PTPRZ inhibits stem cell-like properties and tumorigenicity in glioblastoma cells. Sci Rep. 2017 Jul 17;7(1):5609.

12. Xie Y, Ding W, Xiang Y, Wang X, Yang J. Calponin 3 Acts as a Potential Diagnostic and Prognostic Marker and Promotes Glioma Cell Proliferation, Migration, and Invasion. World Neurosurg. 2022 Sep;165:e721–e731.

13. Mehrian-Shai R, Yalon M, Simon AJ, Eyal E, Pismenyuk T, Moshe I, Constantini S, Toren A. High metallothionein predicts poor survival in glioblastoma multiforme. BMC Med Genomics. 2015 Oct 22;8:68.

14. Makowska M, Smolarz B, Romanowicz H. microRNAs (miRNAs) in Glioblastoma Multiforme (GBM)-Recent Literature Review. Int J Mol Sci. 2023 Feb 9;24(4):3521.

15. Xu H, Zong H, Ma C, Ming X, Shang M, Li K, He X, Du H, Cao L. Epidermal growth factor receptor in glioblastoma. Oncol Lett. 2017 Jul;14(1):512–516.

16. Zhang L, Dimberg A. Pleiotrophin is a driver of vascular abnormalization in glioblastoma. Mol Cell Oncol. 2016 Feb 18;3(6):e1141087.

17. Liu Y, Xu X, Zhang Y, Mo Y, Sun X, Shu L, Ke Y. Paradoxical role of β8 integrin on angiogenesis and vasculogenic mimicry in glioblastoma. Cell Death Dis. 2022 Jun 8;13(6):536.

18. Laks DR, Oses-Prieto JA, Alvarado AG, Nakashima J, Chand S, Azzam DB, Gholkar AA, Sperry J, Ludwig K, Condro MC, Nazarian S, Cardenas A, Shih MYS, Damoiseaux R, France B, Orozco N, Visnyei K, Crisman TJ, Gao F, Torres JZ, Coppola G, Burlingame AL, Kornblum HI. A molecular cascade modulates MAP1B and confers resistance to mTOR inhibition in human glioblastoma. Neuro Oncol. 2018 May 18;20(6):764–775.

19. Li S, Dai Z, Yang D, Li W, Dai H, Sun B, Liu X, Xie X, Xu R, Zhao X. Targeting β2 subunit of Na+/K+-ATPase induces glioblastoma cell apoptosis through elevation of intracellular Ca2. Am J Cancer Res. 2019 Jun 1;9(6):1293–1308.

20. Li X, Zou Z, Ma E, Feng S, Han S. Human Glioma Cells Therapy Using ATRA-Induced Differentiation Method to Promote the Inhibitive Effect of TMZ and CCDP. J Healthc Eng. 2021 Oct 29;2021:6717582.

21. D’Amico AG, Maugeri G, Vanella L, Consoli V, Sorrenti V, Bruno F, Federico C, Fallica AN, Pittalà V, D’Agata V. Novel Acetamide-Based HO-1 Inhibitor Counteracts Glioblastoma Progression by Interfering with the Hypoxic-Angiogenic Pathway. Int J Mol Sci. 2024 May 15;25(10):5389.

22. Liu Y, Fang S, Sun Q, Liu B. Anthelmintic drug ivermectin inhibits angiogenesis, growth and survival of glioblastoma through inducing mitochondrial dysfunction and oxidative stress. Biochem Biophys Res Commun. 2016 Nov 18;480(3):415–421.

23. Seidkhani E, Moradi F, Rustamzadeh A, Simorgh S, Shirvalilou S, Mehdizadeh M, Dehghani H, Akbarnejad Z, Motevalian M, Gorgich EAC. Intranasal delivery of sunitinib: A new therapeutic approach for targeting angiogenesis of glioblastoma. Toxicol Appl Pharmacol. 2023 Dec 15;481:116754.

24. Dhungel L, Rowsey ME, Harris C, Raucher D. Synergistic Effects of Temozolomide and Doxorubicin in the Treatment of Glioblastoma Multiforme: Enhancing Efficacy through Combination Therapy. Molecules. 2024 Feb 14;29(4):840.

